# Haemovigilance and Trends of Transfusion Transmissible Viral Infections among Asymptomatic Population at Akatsi South Municipal in Volta Region of Ghana from 2014 to 2019

**DOI:** 10.1101/2022.12.25.22283933

**Authors:** Abdul-Wahab Mawuko Hamid, Moses Oduro-Mensah, Ishmael Adase, Precious Kwablah Kwadzokpui, Kenneth Owusu Agyemang, Pascal Ayivor, Kofi Karikari Bonsu, Salifu Nanga, Ahmed Tijani Bawah, Huseini Wiisibie Alidu, Israel Tordzro Agudze, Nathaniel Glover-Meni, Ibrahim Jamfaru, Robert Kaba, Ali Mahmudu Ayamba, Theophilus Benjamin Kwofie, Theophilus Adiku, Eric Kwasi Ofori

**Affiliations:** Department of Medical Laboratory Sciences, School of Health and Allied Sciences, University of Health and Allied Sciences, PMB31, Ho Volta Region, Ghana; Department of Clinical Laboratory, Ho Teaching Hospital, PMB 31, Ho, Volta Region, Ghana; Akatsi Municipal Hospital, Agbalixorme on Tadzewu-Akatsi RD, Box Ak83.Akatsi, Volta Region, Ghana; Akatsi Municipal Hospital, Agbalixorme on Tadzewu-Akatsi RD, Box Ak83. Akatsi, Volta Region, Ghana; Department of Basic Sciences, School of Basic and Biomedical Sciences, University of Health and Allied Sciences, PMB 31, Ho, Volta Region, Ghana; Faculty of Allied Health and Pharmaceutical SciencesTamale Technical University, P.O. Box 3E/R, Tamale, Ghana; Department of Medical Laboratory Sciences, School of Allied Health Sciences, University of Health and Allied Sciences, PMB 31, Ho, Volta Region, Ghana; Directorate, School of Allied Health Sciences, University of Health and Allied Sciences, PMB 31, Ho, Volta Region, Ghana; Department of General and Libral Study, School of Basic and Biomedical Sciences, University of Health and Allied Sciences, PMB 31, Ho, Volta Region, Ghana; Department of Microbiology & Immunology, School of Medicine, University of Health and Allied Sciences, PMB 31, Ho, Volta Region, Ghana; Institute of Health Research, University of Health and Allied Sciences, PMB 31, Ho, Volta Region, Ghana; Department of Surgery, School of Medicine, University of Health and Allied Sciences, PMB 31, Ho, Volta Region, Ghana

**Keywords:** Haemovigilance, Transfusion Transmissible Infections (TTIs), Human Immunodeficiency Virus (HIV), Hepatitis Viruses

## Abstract

**Background:** Tracking the changing epidemiology of Transfusion Transmissible Infections (TTIs), including Immunodeficiency virus (HIV), is critical to attaining the Sustainable Development Goals (SDG.3.3) milestones and deadlines. This study assessed the dynamics associated with Blood Donation and TTIs among blood donors at the Akatsi South Municipality in the Volta Region of Ghana.

**Methods:** This was a haemovigilance study, designed to retrospectively evaluate secondary data on 2,588 blood donors in Akatsi South District Hospital from 2014 to 2019. Data was collected, managed and quality controlled done electronically using Microsoft Visual Basics, and STATA. TTIs’ trends were determined using frequentist and descriptive statistics, and 95% confidence intervals using Clopper Pearson test.

**Results:** Prevalence of TTIs was 8.0%. The prevalence of HIV as well as HBV and HCV, was 3.8%, 3.2% and 1.0% respectively. For female hosts, the prevalence was 7.4% (HIV), 4.2 % (HBV) and 1.6% (HCV). For Male-host, the rates were 3.1% (HIV), 3.5% (HBV) and 1.0% (HVC). Donors aged 15-19years were most infected at rates of 13.2% (HIV), 4.7% (HBV) and 1.9% (HCV).

**Conclusion:** About 57(2.4%) and 3(1.2%) of 2380 blood donated were TTIs false negatives and false positives respectively. In addition to being a driver of TTIs among blood donors in this study, the HIV prevalence among teenagers was significantly above the regional and national rates. These rates have ‘programmatic’ and ‘research’ implications. A relatively higher sensitive blood transfusion screening method is urgently needed to prevent the transfusion of TTIs false negative bloods in Akatsi Municipal Hospital

- **What is already known about the topic**: National and global epidemiology of TTIs, including HIV are known.
- **What this study add?** This study serves as baseline data on trends of HIV, HBV and HCV infection among non-sentinel asymptomatic population at Akatsi South Municipality in Volta Region of Ghana.
- **How this study might affect research, practice or policy?** Our data shall contribute to understanding on changing epidemiology of Transfusion Transmissible viral infection including HIV after the introduction of the public health sector strategy toward the 2030 deadline to achieve the Sustainable Development Goal 3.3.

## 1. Introduction

The limited availability of cutting edge technologies for optimum Blood banking services in developing countries including Ghana is a known challenge that compromises the public health strategy to prevent the transfusion of false negative transmissible infections including HIV(1–3). The international response to mitigate risks associated with Blood transfusion has been the recommendations that state actors must initiate centralized national policy and programmes (1,4), ensuring their linkages to “Sustainable Development Goals” (2) and “Blood Safety Information System” (1,3).

Being a subscriber to these conventions, the Ghana Blood transfusion service implemented a “national blood policy, which is driven by “legislative framework”, and it aimed at sustaining a safe blood supply through voluntary blood donation mechanism nationally (5). Consequently, Ghana became the second after Lesotho to launch a “Blood Safety Information System” (BSIS) (3). This project is designed to link HIV infected donors to treatment mechanism. By act of implementation, the use of BSIS tool is projected to transcend from national to regional and district levels in Ghana (3). With less than a decade to meet the SDGs deadline, it is essential to evaluate records on Blood donation so as to track any changing epidemiology of TTIs at the district levels after the implementation of the global health sector strategy in 2016 towards the 2020 milestone and eventually, 2030 deadline. Therefore, this article presents an assessment on the Sero-epidemiologic features of Transfusion Transmissible Infections, including HIV, HBV and HCV among population of blood donors at Akatsi South Municipality in Volta Region of Ghana.

## 2. Materials and Methods

### 2.1 Study Design

This is a haemovigilance study, designed to retrospectively evaluate secondary data on 2, 588 blood donors registered from January 2014 to December 2019 at the Blood bank.

### 2.2 Study Sitting

The study was conducted at Akatsi South Municipal Hospital (Figure1) (6). The Municipality is one of the eighteen municipalities and districts in Volta Region of Ghana (6). It is located in the south-eastern part of the Volta Region at coordinates: 6°S-7°N, 0°W 1°E. The total land area is about 531km^2^. The population was “92, 494 with 43,062 males and 49,432 females” (6). The hospital is a 70-bed capacity facility, which provides a wider range of clinical services, including surgical, reproductive child health and maternity care (6). It serves as a referral center for the surrounding primary health facilities including 4 health centers, 26 National Community Health Planning and Services (CHPS) compounds, and 4 private health facilities in the municipality. As at 2020, the staff strength was over 200.These include 1 Biomedical Scientist and 3 laboratory assistants, who were the technical handlers of Blood banking in the hospital (7).

**Figure 1.**
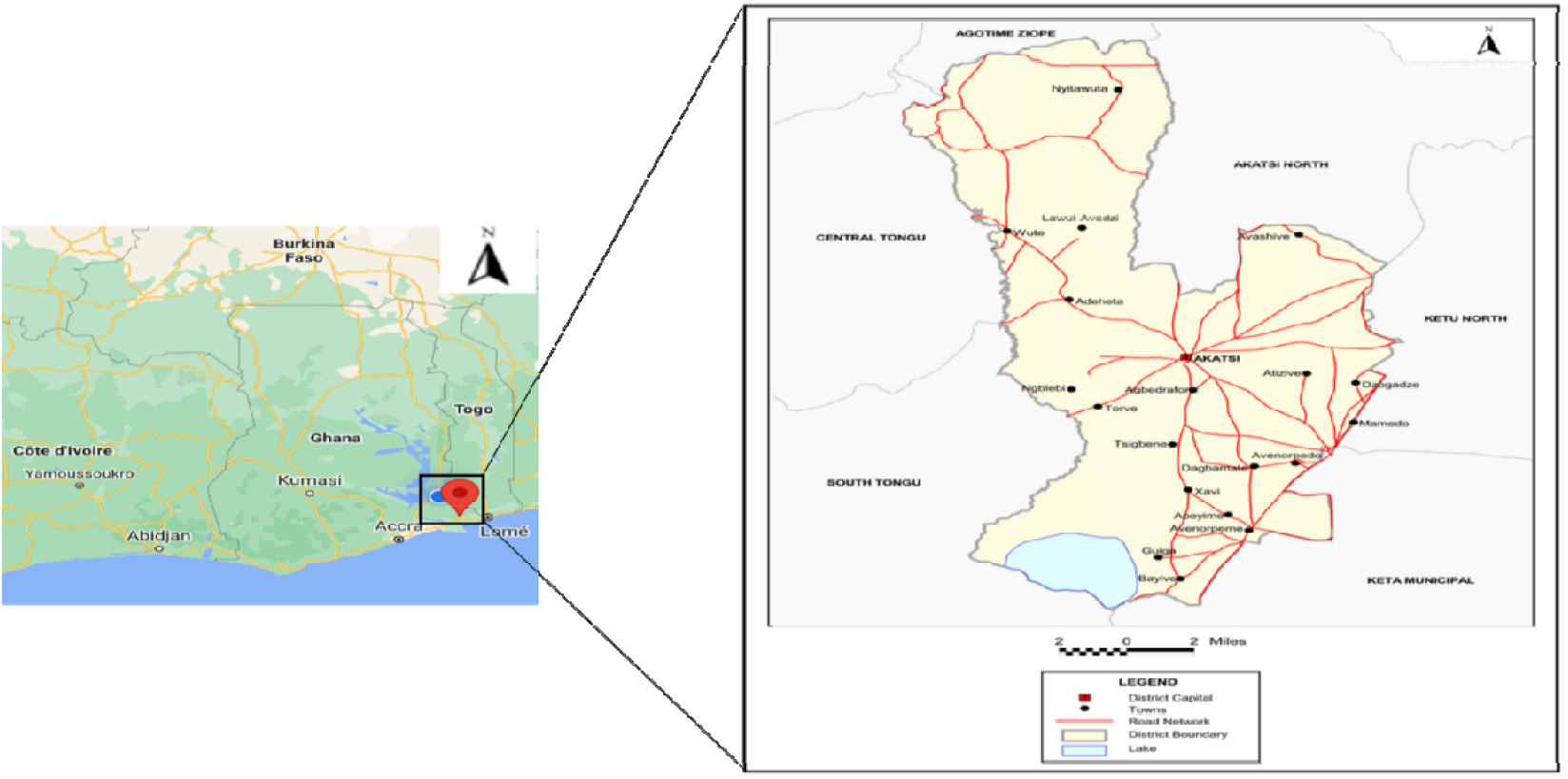
*District Map of Akatsi* South: (6)

### 2.3 Study participants

Prospective blood donors were screened for HIV, HBV and HCV infections using Immunochromatographic Rapid Diagnostic Test kits of about 5 different brands listed in Table 1 over the study period. All the tests were conducted according to the manufacturers’ instructions outlined in the leaflets inserted in the test kits.

**Table 1:** Brands and Validity values of RDT test devices used for screening of HBV, HCV and HIV among Blood donors in Akatsi municipal Blood bank 2014 - 2019

| *Manufacturers* | *Year used* | *Device type* | *Sensitivity* | *Specificity* |
| --- | --- | --- | --- | --- |
| 1. DiaSpot®, USA | 2013-2014 | kit | 99% | 97% |
| 2. ACON, San Diego, CA | 2015 | Strip | 93% | 100% |
| 3. ACON, San Diego, CA | 2016 | Strip | 99% | 99% |
| 4. OneStep HIGHTOP | 2017-2019 | Strip | 99.3% | 99.2% |
| **Average** |  |  | **97.6** | **98.8** |

### 2.4 Data collection, Management and Analysis

The data on socio-demographic and clinical outcomes were collected using extraction log. The data was quality controlled using double data entry mechanism, managed electronically using Microsoft Visual basic, and cleaned data was exported onto STATA software for statistical analyses. The frequentist, Cochrane-Armitage and Clopper Pearson statistics were used to ascertain trends of TTIs.

### 2.5 Ethical issues

Ethical clearance was sought from the University of Health and Allied Sciences, Research Ethics Committee (REC). The certification number is UHAS-REC A.6 [102] 19-20. Permission was also obtained from the Akatsi South Municipal Hospital before the commencement of the study.

## 3. Results

Out of the 2588 participants, 2348 (90.7%) passed, while 240 (9.3%) failed the screening mechanism. Of 240 that failed, 208 (86.7%) and 32 (13.3%) were due to TTIs and clinical limitations respectively. There was no significant variation in proportions of TTIs among voluntary (7.6%) and replacement (8.4%) donors (*x*^*2*^=0.5, p=0.4) (Table3). But there was no significant variation in proportions of TTIs among first time (8.2%) and multiple times (3.3) (*x*^*2*^=2.9, p=0.08) (Table3).

Although, the proportion of female donors (7.3%) were significantly lower than their male counterpart (92.7%) (*x*^*2*^=20.1, p<0.001) (Table2), the proportion of HIV infection was significantly higher (7.4%) among the females than the male donors (3.5%) (*x*^*2*^, p<0.01) (Figure1). The relatively higher prevalence of 7.4% HIV infected females was due to high peaks in 2016 and 2018 (Figure3). Descriptively, the average rate of 4.2% HIV infected female trended significantly by 0.3% above the mean of 3.9 for HIV infected females. Although, the rate of 6.6% HIV infected male in 2016 trended significantly by 0.7% above the mean, the rates of HIV infected male in 2014, 2017, 2018, and 2019 oscillated naturally around the mean (Figure1).

**Table 2:** General Overview of Haemovigilance outcomes and characteristics of study participants

| PARAMETERS | TOTAL | SCREENING OUTCOME | | Cochrane's<br>Mantel-Haenszel<br>Chi-Squire<br>$X^2(p\text{-value})$ |
| --- | --- | --- | --- | --- |
|  |  | *Passed* | *Failed* |  |
|  | *n (%)* | *n (%) [95% CI]* | *n (%) [95% CI]* |  |
| **1. Blood Donors screened:** | 2588 | 2348(90.7)[89.5-91.8] | 240(9.3)[8.2-10.5] | 20.1(<0.01) |
| **2. Blood Sources** |  |  |  |  |
| a) *Voluntary Donation* | 1193(46.1) | 1092(91.5) [89.8-93.1] | 101(8.5) [6.9-10.2] | 1.72(0.19) |
| b) *Replacement Donation* | 1395(53.9) | 1256(90.0) [88.3-91.6] | 139(10.0) [8.4-11.7] |  |
| **3. Donation History** |  |  |  |  |
| a) *First time donors* | 2496(96.4) | 2260(90.5) [89.3-91.7] | 236(9.5) [8.3-10.7] | 2.75(0.09) |
| b) *Multiple donors* | 92(3.6) | 88(95.7) [89.2-98.8] | 4(4.3) [1.2-10.8] |  |
| **4. Demographics** |  |  |  |  |
| a) **Sex** |  |  |  |  |
| i. *Male* | 2398(92.7) | 2188(91.2) [90.0-92.3] | 210(8.8) [7.7-10.0] | 10.3(<0.01) |
| ii. *Female* | 190(7.3) | 160(84.2) [78.2-89.1] | 30(15.8) [10.9-21.8] |  |
| b) **Age Group** |  |  |  |  |
| iii. *15-19* | 190(7.3) | 156(82.1) [75.9-87.3] | 34(17.9) [12.7-24.1] | 1.3(0.25)** |
| iv. *20-29* | 1449(56.0) | 1331(91.9) [90.3-93.2] | 118(8.1) [6.8-9.7] |  |
| v. *30-39* | 695(26.9) | 631(90.8) [88.4-92.8] | 64(9.2) [7.2-11.6] |  |
| vi. *40-49* | 201(7.8) | 184(91.5) [86.8-95.0] | 17(8.5) [5.0-13.2] |  |
| vii. *≥50* | 53(2.0) | 46(86.8) [74.7-94.5] | 7(13.2) [5.5-25.3] |  |

Key. \*\* Cochrane-Armitage Chi-square for trend; CI= Confidence Intervals. $X^2$ =Chi-Squire

**Figure 1:**
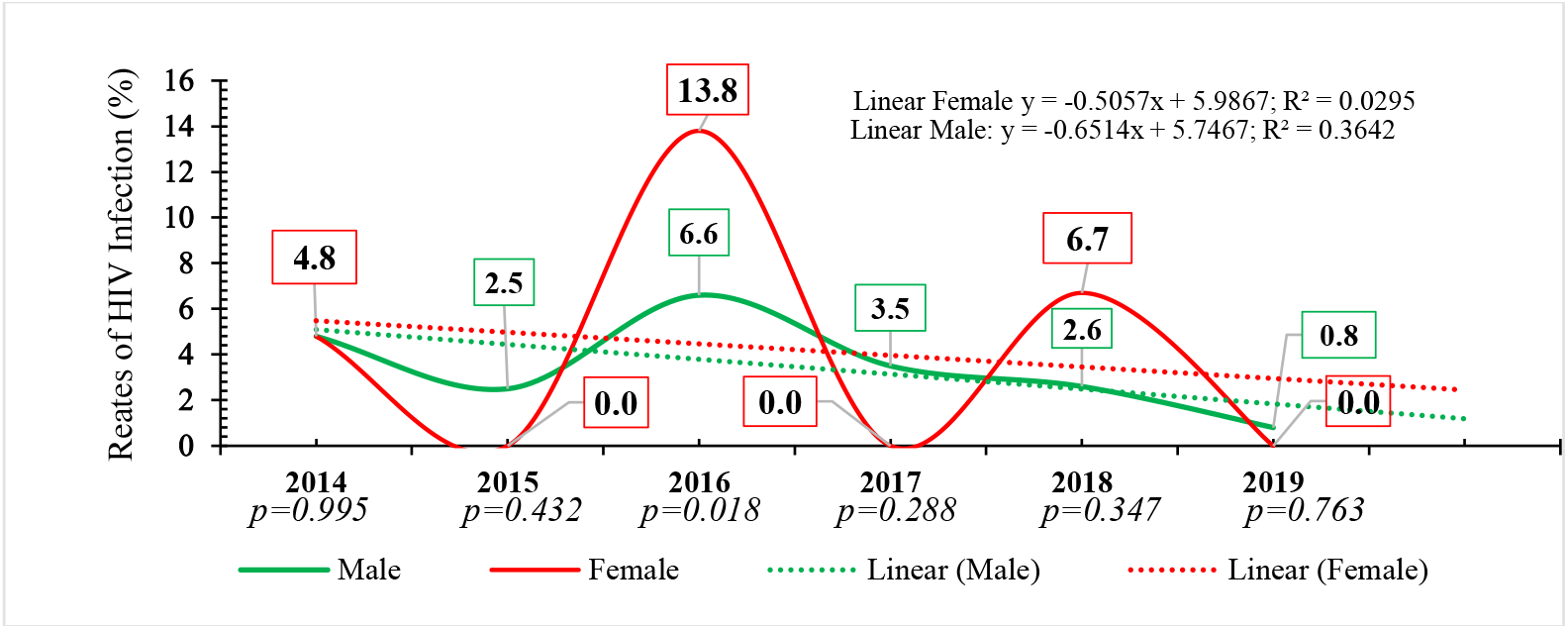
HIV prevalence; Male=3.5%; Female=7.4%; p<0.01 Trends of HIV infection by Host-Gender and Periodic interactions among blood Donors in Akatsi South Municipal Hospital, 2014-202019

**Figure 2:**
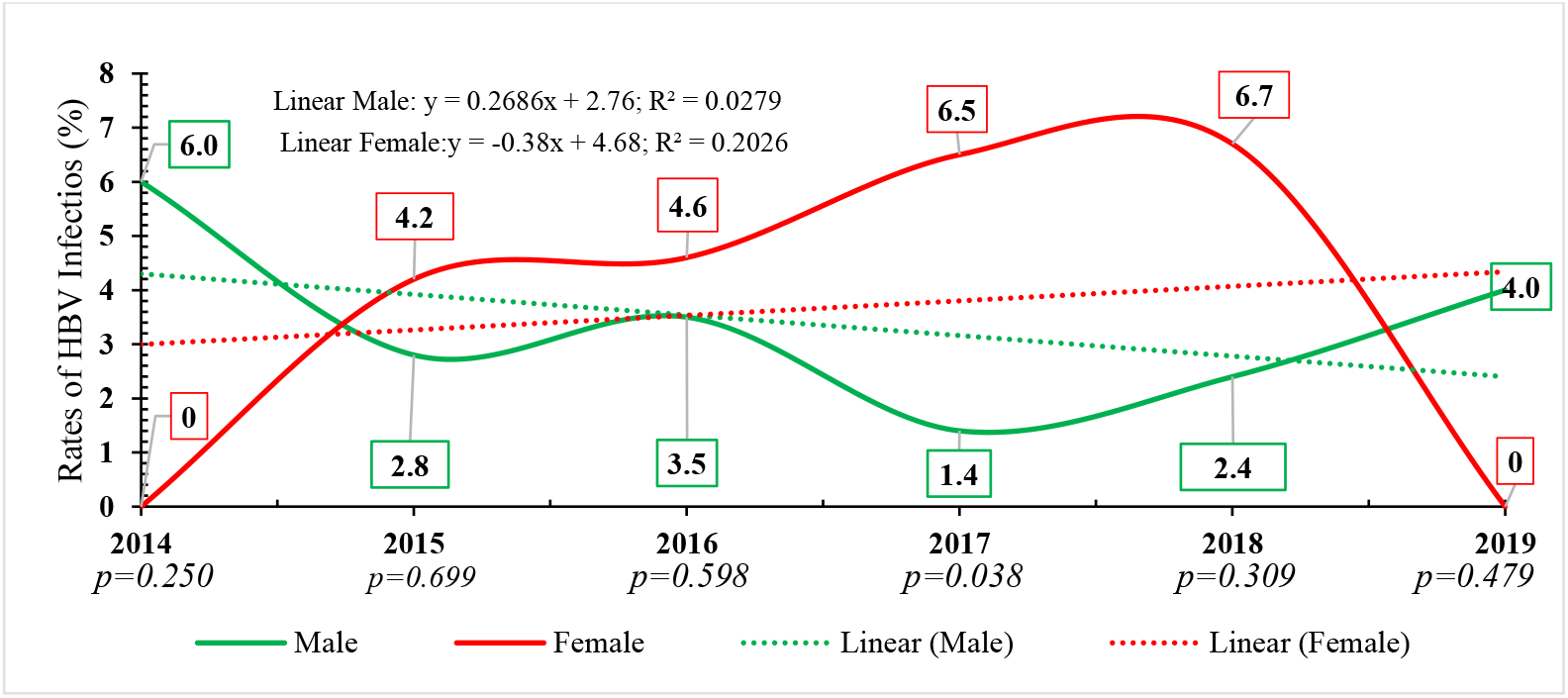
HBV prevalence: Male=3.1%; Female=4.3%; X^2^,P=0.394 Trends of HBV infection by Host-Gender and Periodic interactions among blood Donors in Akatsi South Municipal Hospital, 2014-202019

**Figure 3:**
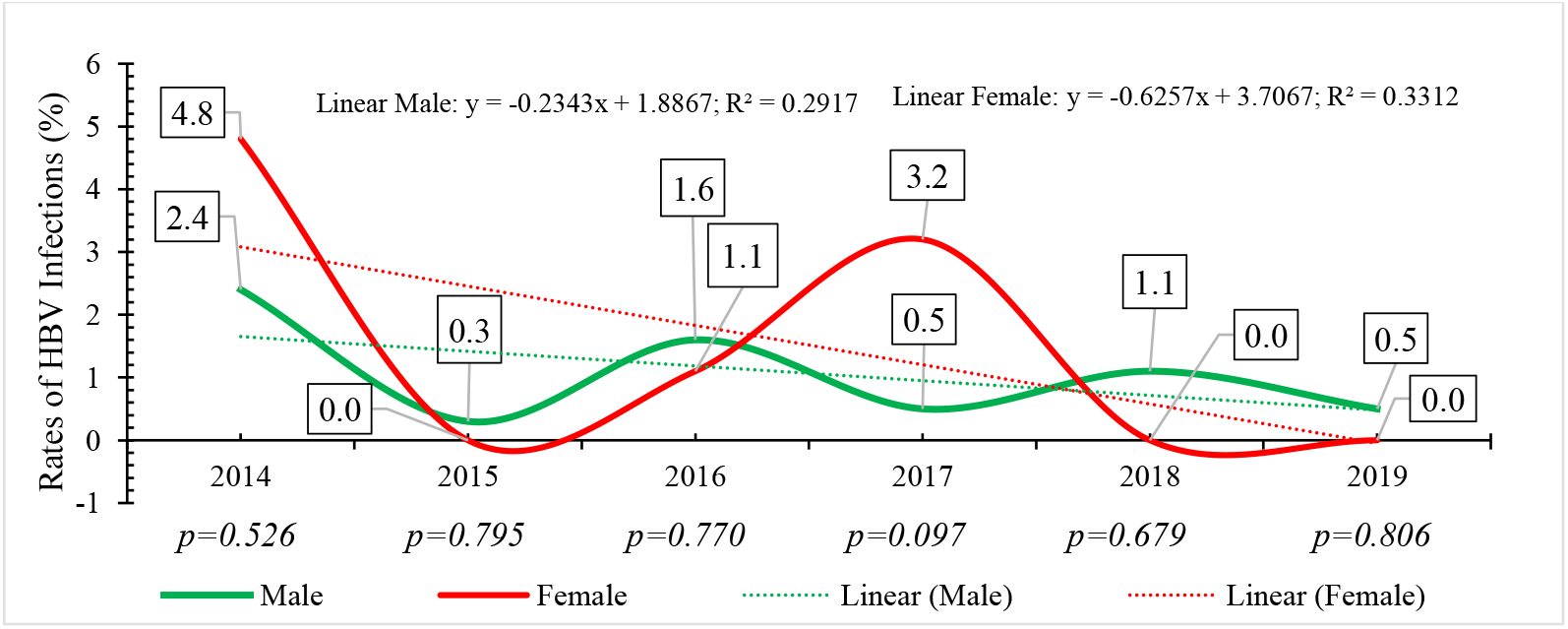
HCV prevalence; Male=1.0%; Female=1.6%; P=0.450 Trends of HCV infection by Host-Gender and Periodic interactions among blood Donors in Akatsi South Municipal Hospital, 2014-202019

Unlike HIV, there was no significant variation in proportions of HBV between female (4.7%) and male (3.1%) (*x*^*2*^=20.1, p=0.39). But the enter variants trends of HBV between female and male were strongly negative correlated (r=-0.9). Similar to HBV, there was no significant variation in proportions of HCV infected female (1.6%) and male (1.0%). Also, the enter trends of HCV infections between female and male were moderately negative correlated (r=-0.6).

The crude prevalence of TTIs was 8.0% (208 of 2588). The point prevalence of TTIs across the period correlated negatively to year-to-year distributions (r=-0.8). Specifically, the highest point prevalence of 12.8% TTIs was in 2014, which was followed by 12.7% (2016), 6.4% (2018), 5.8% (2017) and 5.1% (2019). The variation in proportions of TTIs across the period is statistically significant (*x*^*2*^=11.5, p<0.001). While the rates of TTIs in 2014 and 2016 trended significantly at an average of 2.6% above a mean of 8.1% points, the rates of TTIs in 2015, 2017, 2018 and 2019 distributed naturally at an average rate of 5.7% around the mean. While the trend of TTIs in 2014 was driven by a relatively higher rate of HBV in 2014 [5.3% (10 of 188)], the trend in 2016 was driven by a relatively higher rates of HIV in 2016 [7.5% (50 of 664] (Table 3).

**Table 3:**
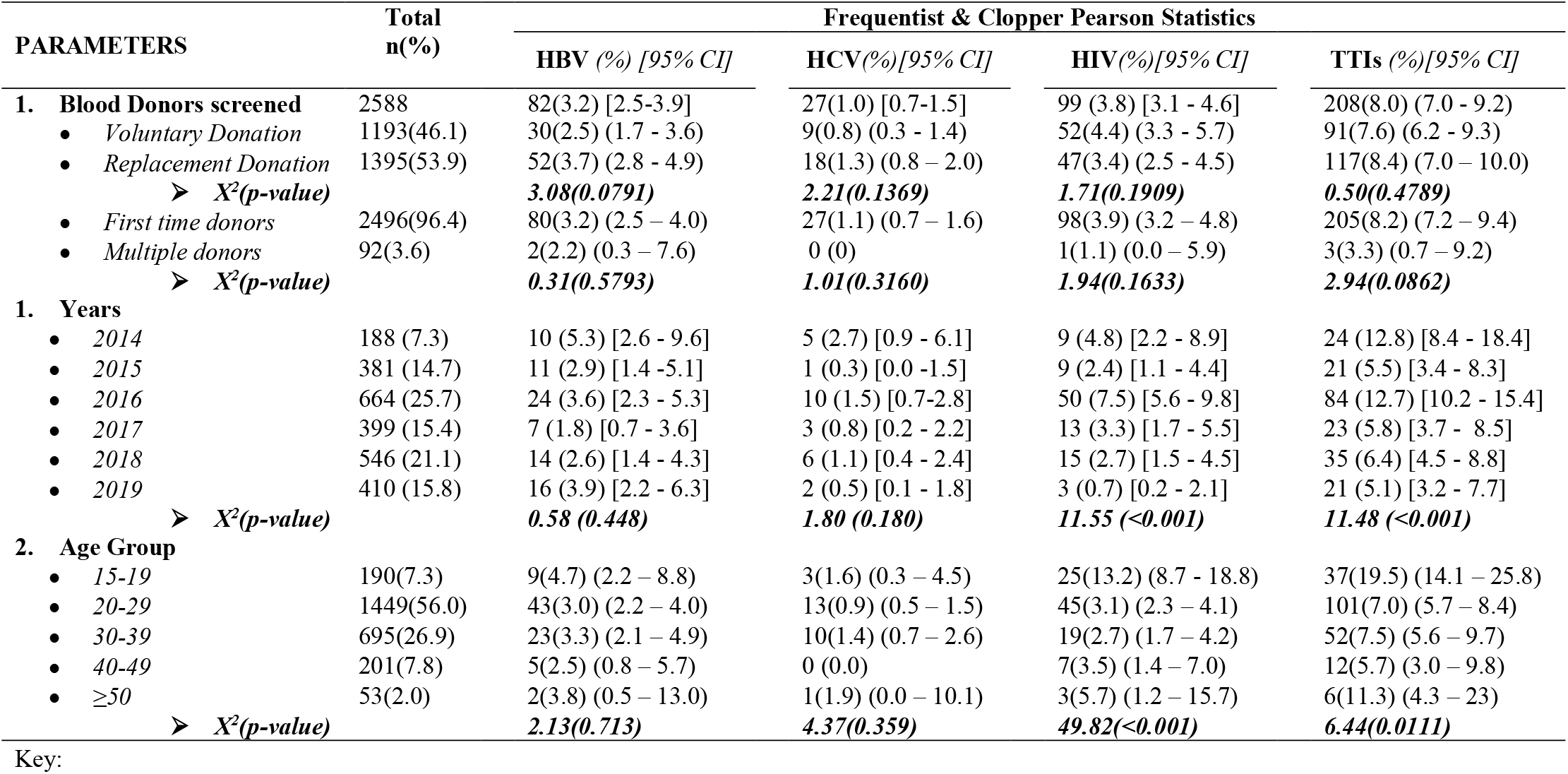
Distribution of TTIs by Donor, Periodic and Socio-demographic parameters in Akatsi Municipal Hospital,2014-2019

| PARAMETERS | Total<br>n(%) | Frequentist & Clopper Pearson Statistics |  |  |  |
| --- | --- | --- | --- | --- | --- |
|  |  | HBV (%) [95% CI] | HCV(%) [95% CI] | HIV(%) [95% CI] | TTIs (%) [95% CI] |
| **1. Blood Donors screened** | 2588 | 82(3.2) [2.5-3.9] | 27(1.0) [0.7-1.5] | 99 (3.8) [3.1 - 4.6] | 208(8.0) (7.0 - 9.2) |
| • Voluntary Donation | 1193(46.1) | 30(2.5) (1.7 - 3.6) | 9(0.8) (0.3 - 1.4) | 52(4.4) (3.3 - 5.7) | 91(7.6) (6.2 - 9.3) |
| • Replacement Donation | 1395(53.9) | 52(3.7) (2.8 - 4.9) | 18(1.3) (0.8 - 2.0) | 47(3.4) (2.5 - 4.5) | 117(8.4) (7.0 - 10.0) |
| ➤ $X^2(p\text{-value})$ | | **3.08(0.0791)** | **2.21(0.1369)** | **1.71(0.1909)** | **0.50(0.4789)** |
| • First time donors | 2496(96.4) | 80(3.2) (2.5 - 4.0) | 27(1.1) (0.7 - 1.6) | 98(3.9) (3.2 - 4.8) | 205(8.2) (7.2 - 9.4) |
| • Multiple donors | 92(3.6) | 2(2.2) (0.3 - 7.6) | 0 (0) | 1(1.1) (0.0 - 5.9) | 3(3.3) (0.7 - 9.2) |
| ➤ $X^2(p\text{-value})$ | | **0.31(0.5793)** | **1.01(0.3160)** | **1.94(0.1633)** | **2.94(0.0862)** |
| **1. Years** |  |  |  |  |  |
| • 2014 | 188 (7.3) | 10 (5.3) [2.6 - 9.6] | 5 (2.7) [0.9 - 6.1] | 9 (4.8) [2.2 - 8.9] | 24 (12.8) [8.4 - 18.4] |
| • 2015 | 381 (14.7) | 11 (2.9) [1.4 - 5.1] | 1 (0.3) [0.0 - 1.5] | 9 (2.4) [1.1 - 4.4] | 21 (5.5) [3.4 - 8.3] |
| • 2016 | 664 (25.7) | 24 (3.6) [2.3 - 5.3] | 10 (1.5) [0.7-2.8] | 50 (7.5) [5.6 - 9.8] | 84 (12.7) [10.2 - 15.4] |
| • 2017 | 399 (15.4) | 7 (1.8) [0.7 - 3.6] | 3 (0.8) [0.2 - 2.2] | 13 (3.3) [1.7 - 5.5] | 23 (5.8) [3.7 - 8.5] |
| • 2018 | 546 (21.1) | 14 (2.6) [1.4 - 4.3] | 6 (1.1) [0.4 - 2.4] | 15 (2.7) [1.5 - 4.5] | 35 (6.4) [4.5 - 8.8] |
| • 2019 | 410 (15.8) | 16 (3.9) [2.2 - 6.3] | 2 (0.5) [0.1 - 1.8] | 3 (0.7) [0.2 - 2.1] | 21 (5.1) [3.2 - 7.7] |
| ➤ $X^2(p\text{-value})$ | | **0.58 (0.448)** | **1.80 (0.180)** | **11.55 (<0.001)** | **11.48 (<0.001)** |
| **2. Age Group** |  |  |  |  |  |
| • 15-19 | 190(7.3) | 9(4.7) (2.2 - 8.8) | 3(1.6) (0.3 - 4.5) | 25(13.2) (8.7 - 18.8) | 37(19.5) (14.1 - 25.8) |
| • 20-29 | 1449(56.0) | 43(3.0) (2.2 - 4.0) | 13(0.9) (0.5 - 1.5) | 45(3.1) (2.3 - 4.1) | 101(7.0) (5.7 - 8.4) |
| • 30-39 | 695(26.9) | 23(3.3) (2.1 - 4.9) | 10(1.4) (0.7 - 2.6) | 19(2.7) (1.7 - 4.2) | 52(7.5) (5.6 - 9.7) |
| • 40-49 | 201(7.8) | 5(2.5) (0.8 - 5.7) | 0 (0.0) | 7(3.5) (1.4 - 7.0) | 12(5.7) (3.0 - 9.8) |
| • ≥50 | 53(2.0) | 2(3.8) (0.5 - 13.0) | 1(1.9) (0.0 - 10.1) | 3(5.7) (1.2 - 15.7) | 6(11.3) (4.3 - 23) |
| ➤ $X^2(p\text{-value})$ | | **2.13(0.713)** | **4.37(0.359)** | **49.82(<0.001)** | **6.44(0.0111)** |

In addition, neither the frequentist showed a significant variation in proportion of donor-age groups; nor the Cochrane-Armitage showed a significant trend of TTIs by donor-age around the central theorem (*x*^*2*^=1.3, p=0.25). The highest rate of TTIs (19.5%) was among donors’ age ≤19 years; followed by age ≥50 (11.3%; 6 of 53), age 30-39 (7.5%; 52 of 695), age 20-29 (7.0%; 101 of 1449) and age 40 – 49 (5.7%; 12 of 201). The variation in proportions of TTIs by donor-age distribution was statistically significant (*x*^*2*^=6.4, p=0.01). The TTIs rates among ≤19years trended significantly by 7.3% above a mean of 10.2, and the TTIs among ≥50 years trended significantly by 1.1% above the mean point. But the TTIs rates among age 20-49 oscillated naturally at a rate of 6.7% around a mean point. While the trend of TTIs among age ≤19 was driven by a relatively higher rate of HIV infection [13.2% (25 of 190)], the TTIs’ trend among age ≥50 was also driven by a relatively higher rate of HIV infections [5.7% (2 of 53)] (Table3).

## 5. Discussion

In terms of blood transfusion services, the trends of blood donor population was dynamic, and observed intermitted troughs in 2014, 2015, 2017 and 2019, and intermitted peaks in 2016 and 2018 could be a reflection of limitations associated with blood banking to sustain the blood mobilization strategies (8,9). Also, the preponderance of male over female donors was also reported in number of studies across the regions in Ghana (10,11), as well as in sub-Saharan Africa (12,13) and Persian Gulf (14,15). The most cited reasons for this phenomenal low female donors included exhaustive lists of socio-cultural beliefs and clinical parameters such as menstruation cycles and maternity (8,10,11).

The relatively higher proportion of donors age 20-29 (56.0%) was similar to studies in Ethiopia (12,13). The reasons attributed for these development included the dependency on secondary school and college students to sustain the blood banks (8,9). This effort to use students, plus the fact that the population pyramid in Akatsi South Municipality was characterized by a relatively higher proportion of youths (6), could have driven the dominance of younger aged blood donors in our study. Expectedly, the greater proportions of 96.4% blood sources were first time donors, and most of them (53.9%) were for family replacement (16). Globally, because of the low repeats of donors as observed in this study (3.6%), blood banks often have to rely on family replacement donors; most of whom are never prepared for blood donation (1,16,17).

Moreover, in respect of trends of the TTIs, the 8.0% rate of TTIs in this study was within the range of 3.6% in Ho (18) and 18.3% in Hohoe in the study region (19). A comparable higher rate of 19.5% TTIs was reported in the middle zone of Ghana (20). Nonetheless, our TTIs rate was significantly higher than the 0.3% and 2.4% reported in Persian (21) and sub-Saharan Africa (22). The high rate of TTIs in this study was driven by the high pool of replacement donors. Indeed, it is this risks of TTIs associated with replacement donors that WHO recommends the adherence to voluntary donations (23). Furthermore, the variations in burden of TTIs across the aforementioned studies could be a reflection of natural variations associated with geo-demographic endemicity of venereal infections across the regions (19). A stratified analysis on donor-age depicted a very high prevalence of TTIs among younger aged-donors. Similar trends was reported in Northern Ghana, where the highest rates of 4.7% HBV and13.2% HIV were observed among a younger aged group 15-19 (24). These developments could be due to limited exposure to sexual education, as well as exposure to drug elicitation among the younger generation(25). Similar to study in Ghana (19), but in contrast to studies in Ethiopia (12,13,22), the named infections were more prevalent among female than male donors in our study. The relatively higher rates of TTIs among female in this study could be due to the natural anatomic limitations associated with female sexual organs, which often compromise the innate defense system (26). Also, the socio-economic exuberantions associated with female population within the municipality and across the border township in Ghana and Togo could have exposed our female donors to higher risks environment and partners(27,28).

Additionally, findings on the transfusion transmissible HIV, suggest the rate of 3.8% HIV infection in this study was lower than 4.8% reported among blood donors in Ho within the study region (29), 4.9% at Kintampo in Bono East region (20). Nevertheless, this rate was significantly higher than the general population based regional and national rates of 1.28% and 2.0% respectively (30). Although the higher proportion of HIV infected female is not unique to this study, the persistence of HIV across the period aligns with the observation made by the locally based HIV/AIDS response team’s assessment on the surge of HIV/AIDS in the municipality (31). Nonetheless, using 2015 as the baseline, a relatively point prevalence of 0.7% HIV (3 of 410) in 2019 was a remarkable reduction of HIV to a rate of 33.3% from 2015 to 2019. This was just a 0.3% higher than the 30% reduction targeted for 2020 by global health strategy towards to ending epidemic HIV/AIDS by 2030 (32).

Likewise, it was observed that the rate of 3.2% in this study was lower than the 6.9% in Ho Municipal (33), and 9.6% in Soboba District in Northern Region (24). Compared to the figures at national level, this rate is significantly below the 12.3% recorded within, MDGs era (34), and 11.3% within the SDGs era (35) in Ghana. The intra low rate of HBV in this study could be due to the impact of the implementation of as well as global policy and national programmes towards the eradication of TTIs, including HIV in Ghana (36). The periodicals depicted a higher mean rate of 4.4% HBV from 2014 to 2015, and a lower mean of 2.9 HBV from 2016 to 2019 in this study. The inter variations in rate of HBV infection could be due to the different testing methods employed in the diagnosis of the viral infections (37) and/ or a reflection of natural ecological variations between the study sites(38). The highest rate of 4.7% HBV among younger age donors (≤19), and rate of 4.9% HBV among older donors (≥50) was also reported in Northern Ghana (24). However, the variation between rate of new infection (younger aged) and chronic carriers (older aged) was insignificantly by 0.2% point. This implicates the public health interventions to reduce the margins of new TTIs, including HIV (39)

The rate 1.0% HCV in this study was significantly below the national rate of 3.0% in Ghana (40). The lower rate of HCV affirmed that significant intra and inter variation in HCV prevalence and endemicity exists among different geo-demographics in Ghana. Although, the 1% HCV was a lower rate, the inconsistent pattern in which HCV oscillates among female donors compared to more stable distribution of HCV among male donor across the period is of significant public health concern that requires further research.

### 5.1 Limitations

Although, all attempt were made to minimize the type 1 and type2 errors associated with retrospective study such as this, we were unable to overcome the inherent limitations associated with using Rapid Diagnostic Test (RDT) kits results obtained under non-experimental conditions. Therefore, the findings of our study should be interpreted with caution.

### 5.2 Conclusion

The sero-prevalence of TTIs as exhibited in the study shows significant reductive trends from 2015 to 2019. This was a positive implication on existing public health strategies to combat TTIs, including HIV/AIDS in Akatsi south municipality and Ghana. Despite this positive assessment, the relatedly higher rates of TTIs among female donors who constituted only 7.3% donor population have programmatic and research implications.

### 5.3 Recommendations

First, in order to sustain the reductive trend of named TTIs in this area, it is highly recommendable that the decentralization programmes component of the “National Strategic framework1 and 2” policies on venereal infections are accelerated at the sub-district levels in Ghana. Secondly, the use of chromatographic method (RDTs), with limited validity metrics, is a potential medium for transfusion false negative blood. About 2.4% of the blood transfused within the period were false negatives for the named TTIs (Table1). Therefore, a relatively higher sensitive screening methods such Enzyme Linked Immunosorbent Assay (ELISA) and/or Polymerase Chain Reaction (PCR) methods are urgently needed to prevent the transfusion of TTIs false negative bloods in Akatsi Municipal Hospital.

## Data Availability

All data produced in the present study are available upon reasonable request to the authors

## Notes

### Competing Interest Statement

The authors have declared no competing interest.

### Funding Statement

This study did not receive any funding

### Author Declarations

The University of Health and Allied Sciences' Review Ethical Committee gave ethical approval for this work on certification number UHAS-REC A.6 [102] 19-20.

